# Blood pressure checkups practices and associated factors among federal ministry civil servants, Addis Ababa, Ethiopia, 2024

**DOI:** 10.1101/2025.08.01.25332628

**Authors:** Nibretie Chalachew Anteneh, Tadesse Shiferaw Chekol, Zewdu Minwuyelet Gebremariam

## Abstract

**Background:** Hypertension is a global public health concern, contributing significantly to cardiovascular morbidity and mortality. In Ethiopia, 27.3% of federal ministry civil servants were hypertensive in 2015. Regular blood pressure checkups are vital for early detection and management, yet there is limited data on blood pressure checkup practices in workplace setting, including federal ministries in Ethiopia. Understanding these practices and their determinants is essential for designing targeted work place health interventions.

**Objective:** This study assessed the prevalence of blood pressure checkup practices and associated factors among Federal Ministry civil servants in Addis Ababa, Ethiopia.

**Methods:** A cross-sectional study was conducted among 484 federal ministries civil servants in Addis Ababa, Ethiopia. A simple random sampling technique with proportional allocation was used to select study participants. Data were collected using interviewer administered structured questionnaire. Both bi-variable & multi-variable logistic regression models were employed to identify factors associated with blood pressure checkup practices. Descriptive and inferential statistics were presented with tables and graphs.

**Results:** Among civil servants in federal ministries, 43.8% [95% CI: 39.3–48.4%] reported having checked their blood pressure. Blood pressure checkup practice was significantly associated with age ≥40 years [AOR = 2.95, 95% CI: 1.53–5.70], male sex [AOR = 1.99, 95% CI: 1.25–3.21], good knowledge [AOR = 2.82, 95% CI: 1.73–4.61], positive attitude [AOR = 3.22, 95% CI: 1.99–5.22], and family history of hypertension [AOR = 4.86, 95% CI: 2.47–9.58]. These findings highlight the importance of blood pressure screening among at risk populations in the workplace.

**Conclusion:** Despite the importance of blood pressure checkups, a significant proportion of federal ministry civil servants in Addis Ababa do not regularly check their blood pressure. Interventions targeting knowledge, attitudes, and family history of HTN are necessary to improve blood pressure checkup practices and promote early hypertension management

## Introduction

Hypertension is a leading cause of global morbidity and mortality, particularly in low- and middle-income countries ^[1,2]^. It is a major risk factor for cardiovascular diseases (CVDs) ^[3,4]^. In Ethiopia, the prevalence of hypertension is rising ^[5]^. National data showed a pooled prevalence of 21.8%, with Addis Ababa having the highest rate at 25.4% ^[6]^. Prevalence increases with age from 7.3% (18–39 years) to 65% (≥60 years ^[7]^. A 2015 national survey showed that 60% of hypertensive individuals were unaware of their condition, contributing to 70% of stroke admissions ^[8]^.

Routine medical checkups (RMCs), or periodic health examinations, are crucial for early detection of chronic diseases like hypertension ^[9–14]^. Despite some debate about their cost-effectiveness and potential overdiagnosis ^[15]^, several studies support its role in reducing morbidity and mortality ^[14–16]^. Blood pressure (BP) screening, a core element of RMCs, is essential for timely intervention ^[16]^, but its practice is under-researched in Ethiopia ^[17]^.

The U.S. Preventive Services Task Force and similar global guidelines recommend BP screening starting at age 18 years, with annual checks for high-risk groups and those aged ≥40 years ^[18]^. The Ethiopian Ministry of Health recommends BP screening for adults aged ≥30 years ^[19]^. Despite these recommendations, more than half of hypertensive individuals remain undiagnosed, especially in urbanizing sub-Saharan Africa ^[20,21]^, emphasizing the critical need for regular screening programs ^[22]^.

Globally, studies showed wide variations in BP checkup practice, ranges from 40% in China ^[23]^, 34.3% in Riyadh ^[24]^, 31.7% in Bangladesh ^[4]^, 25.5% in Saudi students ^[25]^ to as low as 4.7% among Myanmar migrants in Thailand ^[26]^. In Ethiopia, blood pressure checkup practice vary from 29.8% among Arba Minch town civil servants ^[17]^ to 8.5% in Kellem Wollega ^[27]^, while the prevalence of hypertension is 27.3% among federal ministry civil servants in Addis Ababa, Ethiopia ^[5]^.

Blood pressure checkup practice is influenced by factors such as age, sex, income, education, knowledge, and attitude ^[17,26, 28–31]^. In Ethiopia, screening rates are higher among older adults (≥60 years), men, and those with higher education ^[21,8,32, 33]^. Family history, physical activity, and workplace program encourage checkups, while smoking, heavy workload, and poor access to services reduce checkup practice ^[17,31,34–39]^. Awareness interventions via media, community health workers, and digital tools can boost BP screening rates ^[40–43]^, while barriers like low health literacy, mistrust, and cultural factors hinder utilization ^[44–49]^.

National strategies and workplace-based screenings are vital for underserved groups ^[50]^. Limited data exist on BP checkup practices among federal civil servants in Addis Ababa ^[5]^. This study aims to fill that gap, offering insights to inform targeted interventions, ultimately improving early detection and reducing hypertension-related complications. To explore factors affecting BP checkup practices among civil servants, a conceptual framework was developed from existing literature ^[17, 26, 28]^ and tailored to the study context. These factors were categorized into five domains: socio-demographics, knowledge, attitude, workplace factors, and other individual-level factors.

## Methods and Materials

### Ethical Statements

Ethical clearance was obtained from the Ethical Review Committee of GAMBY Medical and Business College. A formal letter was written from the college to the respective Federal ministries before going to data collection. The purpose of the research was explained to the eligible participants, what was expected of them, and how long the interviews would last. Participants’ written informed consent indicates that they have the right to withdraw from the study at any time. At all levels of the study, confidentiality was maintained by avoiding the use of names and other identifiers.

### Study design and population

An institution-based cross-sectional study was conducted among civil servants working in federal ministries located in Addis Ababa, Ethiopia, from June 24 to July 30, 2024. Addis Ababa, Ethiopia’s capital and largest urban center, hosts 11 sub-cities, 116 districts, and 22 federal ministries. As 2024 Ethiopian Civil Service Commission report, these ministries collectively employed 18,173 civil servants. The source population included all civil servants working in federal ministries, while the study population included those from a subset of randomly selected ministries at the time of data collection.

### Sample Size Determination and Sampling Technique

A sample size of 508 was calculated using a single population proportion formula based on a 29.8% estimated prevalence of BP checkup practice (from prior research), 95% confidence level, 5% margin of error, 1.5 design effect, and 5% non-response rate. For the second objective, Epi Info was used to calculate sample sizes based on key variables, but the larger sample (508) was retained to ensure adequacy.

A multistage sampling technique was used. First, 7 ministries were randomly selected from the total of 22: the Ministry of Urban Development and Construction, the Ministry of Culture and Sport, Ministry of Education, the Ministry of Health, the Ministry of Transport and Logistics, Ministry of Trade and Regional Integration, and the Ministry of Agriculture. The total sample was proportionally allocated to each selected ministry based on the number of employees.

### Inclusion and Exclusion Criteria

The study included civil servants from selected ministries, while excluding those previously diagnosed with hypertension to avoid bias.

### Data management and Statistical Analysis

Data were collected through an interviewer-administered structured questionnaire covering socio-demographics, knowledge & attitudes, individual health factors, workplace conditions, and BP checkup practices. A pretest was conducted on 5% of the sample at national tissue and blood bank and bole sub city civil servants to ensure clarity. Trained diploma-level health professionals collected the data under daily supervision by the principal investigator to ensure data quality.

Data were entered into Epi-Data version 4.7.0 and analyzed using SPSS version 27. Descriptive statistics were presented using tables and graphs. Binary logistic regression was used to identify factors associated with BP checkup practice, with variables having a p-value <0.25 in bivariable analysis entered into multivariable logistic regression. Adjusted odds ratios (AOR) with 95% confidence intervals and p-values <0.05 were used to determine statistically significant variables.

## Results

The result of Table 1 revealed that a total of 484 federal ministry civil servants participated in the study, with a response rate of 95.3%. The median age of respondents was 40 years (SD ±9.34), with 51.2% (n = 248) aged 40 years and above. Males constituted 57.4% (n=278) of participants, while females made up 42.6% (n =206). Regarding educational attainment, the majority, 78.1% (n = 378), had at least a first degree, whereas 21.9% (n =106) had lower educational levels. In terms of marital status, 73.6% (n= 356) were married, and 26.4% (n=128) were single. Work experience varied, with 65.1% (n = 315) having worked for 10 years or more, and the remaining 34.9% (n =169) having less than 10 years of service. Religiously, the majority identified as Orthodox Christians (60.1%, n= 291), followed by Protestants (24.8%, n= 120), Muslims 13.8%, (n= 67), and others 1.2% (n=6). Monthly income distribution revealed that 36.2% (n=175) earned between 6100–9056 ETB, 25.2% (n=122) earned below 6100 ETB, 18.8% (n=91) earned between 9420–10150 ETB, and 19.8% (n= 96) earned above 10150 ETB.

**Table 1:** Socio-Demographic Characteristics of Respondents Among Federal Ministry Civil Servants in Addis Ababa, Ethiopia.

| Variable | Category | Frequency | Percent |
| --- | --- | --- | --- |
| Age of respondents | < 40 years old | 236 | 48.8 |
|  | ≥ 40 years old | 248 | 51.2 |
| Sex of respondents | Female | 206 | 42.6 |
|  | Male | 278 | 57.4 |
| Level of education | <1<sup>st</sup> Degree | 106 | 21.9 |
|  | ≥ 1<sup>st</sup> Degree | 378 | 78.1 |
| Marital status | Single | 128 | 26.4 |
|  | Married | 356 | 73.6 |
| Work experience | < 10 years | 169 | 34.9 |
|  | ≥10 years | 315 | 65.1 |
| Religion | Orthodox | 291 | 60.1 |
|  | Protestant | 120 | 24.8 |
|  | Muslim | 67 | 13.8 |
|  | Other | 6 | 1.2 |
| Monthly income | < 6100 ETB | 122 | 25.2 |
|  | 6100 – 9056 ETB | 175 | 36.2 |
|  | 9420 – 10150 ETB | 91 | 18.8 |
|  | > 10150 ETB | 96 | 19.8 |

The results of Table 2 showed that study participants had different levels of knowledge and attitudes about BP checkup practice. After categorizing participants’ knowledge and attitude scores, 251 (51.9%) were classified as having good knowledge, and 248(51.20%) were classified as having a positive attitude towards hypertension. Similarly, 72(14.90%) had a family history of HTN, 32(6.60%) had other chronic diseases, 132 (27.30%) performed physical exercise, 10 (2.10%) were current smokers, and 28 (5.80 %) had health insurance.

**Table 2:** Individual factors affecting blood pressure checkup practice of participants among Federal Ministry civil servants in Addis Ababa, Ethiopia.

| Variables | Response Category | Frequency | Percent |
| --- | --- | --- | --- |
| Knowledge of participants | Poor | 233 | 48.1 |
|  | Good | 251 | 51.9 |
| Attitude of participants | Negative | 236 | 48.8 |
|  | Positive | 248 | 51.2 |
| Family history of HTN | No | 412 | 85.1 |
|  | Yes | 72 | 14.9 |
| Other chronic diseases | No | 428 | 88.4 |
|  | Yes | 56 | 11.6 |
| Physical Exercise | No | 316 | 65.3 |
|  | Yes | 168 | 34.7 |
| Currently Smoker | No | 474 | 97.9 |
|  | Yes | 10 | 2.1 |
| Have Health Insurance | No | 438 | 90.5 |
|  | Yes | 46 | 9.5 |

The results of Figure 1 indicated that workplace-related conditions showed a notable influence on blood pressure checkup practices among participants. Of the 484 respondents, 92.6% (n = 448) reported the absence of a workplace blood pressure screening program in their institutions. Only 7.4% (n = 36) had access to such services. Furthermore, 22.9% (n = 111) of participants experienced work overload, while 77.1% (n = 373) reported manageable workloads.

**Figure 1:**
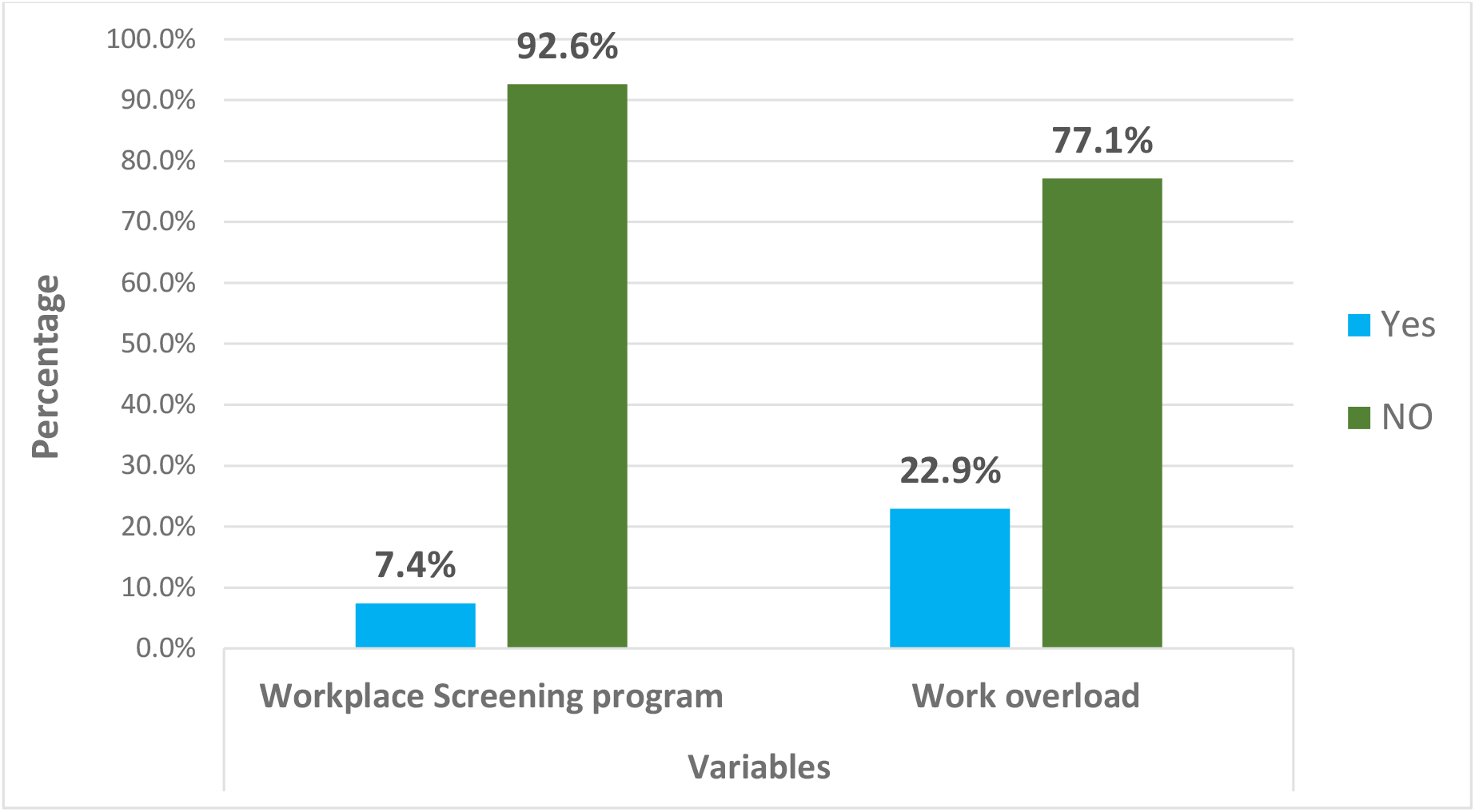
Workplace factors affecting blood pressure checkup practice among federal ministry civil servants, Addis Ababa, Ethiopia.

The results of Figure 2 showed that out of the 484 study participants, 212 (43.80% [95% CI: 39.3% - 48.4%]) checked their blood pressure. Among them, 149 (30.80%) were males, and 63 (13.00%) were females, indicating that males were more likely to have blood pressure checkup practice compared to females.

**Figure 2:**
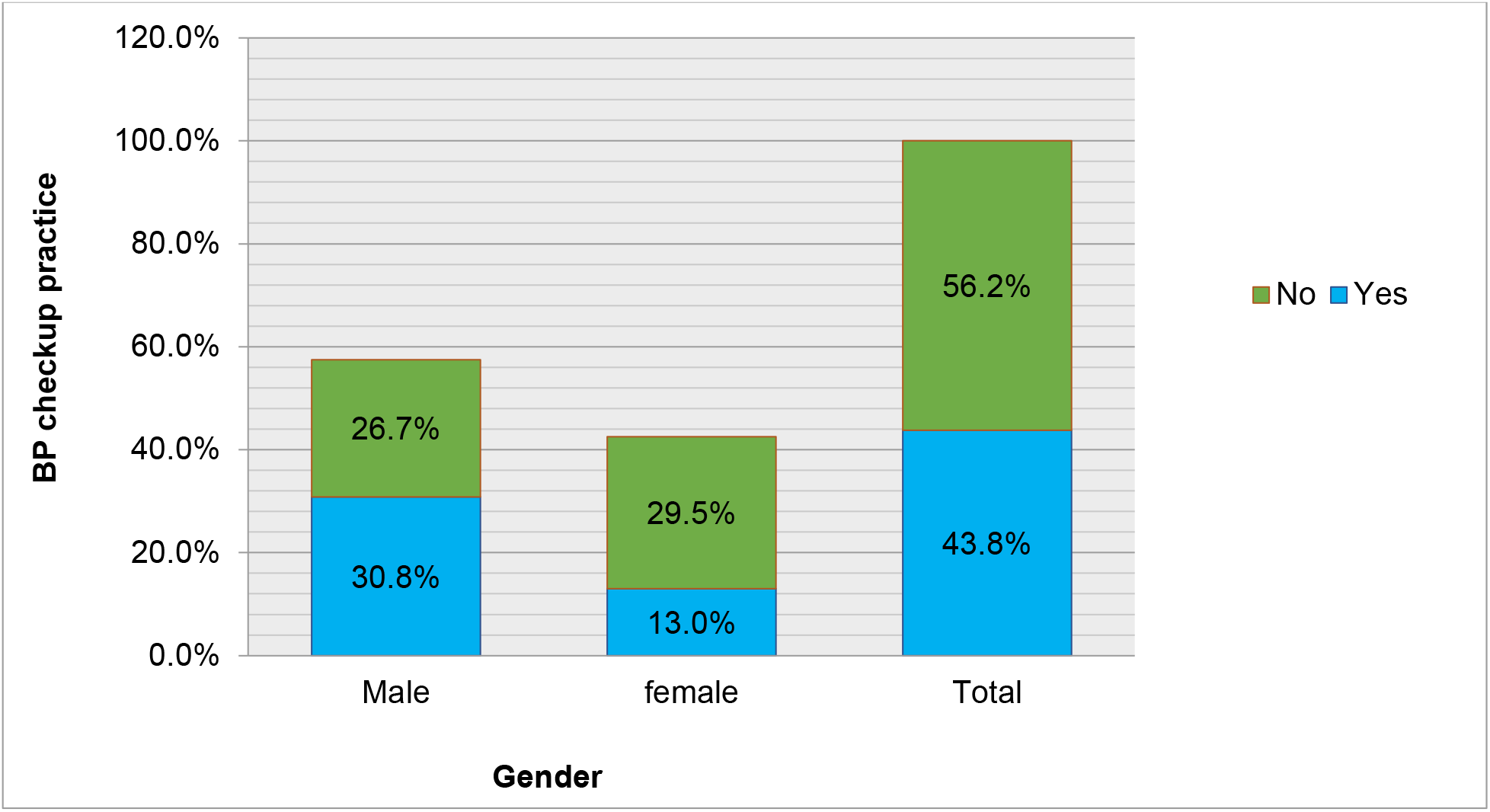
Blood pressure checkup practice among federal ministry civil servants by gender, Addis Ababa, Ethiopia.

In the bivariable logistic regression analysis (**Table 3**), age, sex, level of education, marital status, work experience, knowledge, attitude, and family history of hypertension were significantly associated with blood pressure checkup practice at a p-value threshold of <0.25. However, in the multivariable logistic regression analysis, only five variables remained statistically significant (p < 0.05): age, sex, knowledge, attitude, and family history of hypertension.

**Table 3:** Factors associated with blood pressure checkup practice in the bivariate and multivariate logistic regression analysis among federal ministries’ civil servants in Addis Ababa, Ethiopia, 2024.

| Variables | Category | Frequency | COR [95% C.I] | AOR [95% C.I] |
| --- | --- | --- | --- | --- |
| Age | Age ≥40 years | 247 (51.0%) | 2.79 [1.92, 4.05] * | 2.95 [1.53, 5.70] * |
|  | Age<40 years | 237 (49.0%) | 1 | 1 |
| Sex | Males | 278 (57.4%) | 2.62 [1.80, 3.83] * | 1.99 [1.25, 3.21] * |
|  | Female | 206 (42.6%) | 1 | 1 |
| Level of education | ≥ 1st degree | 378 (78.1%) | 4.44 [2.62, 7.52] | - |
|  | < 1st degree | 106 (21.9%) | 1 | - |
| Marital Status | Married | 175 (36.2%) | 4.05 [2.52, 6.49] | - |
|  | Single | 122 (25.2%) | 1 | - |
| Work experience | ≥ 10 years | 315 (65.1%) | 2.42 [1.63, 3.59] | - |
|  | < 10 years | 169 (34.9%) | 1 | - |
| Knowledge | Good | 248 (51.2%) | 4.99 [3.37, 7.38] * | 2.82 [1.73, 4.61] |
|  | Poor | 233 (48.8%) | 1 | 1 |
| Attitude | Positive | 248 (51.2%) | 5.23 [3.53, 7.76] * | 3.25 [1.98, 5.22] |
|  | Negative | 236 (48.8%) | 1 | 1 |
| Family history of HTN | Yes | 72 (14.9%) | 6.3 [3.45, 11.51] * | 4.86 [2.47, 9.58] |
|  | No | 412 (85.1%) | 0.61 | 1 |

**Note:** 1: Reference, \*= P-value<0.05, COR = Crude odds ratio, AOR = Adjusted odds ratio, and CI = Confidence interval.

Participants aged 40 years and above were nearly three times more likely to check their blood pressure as compared to those under 40 years [AOR = 2.95, 95% CI: 1.53–5.70]. Males were twice more likely to practice BP checkups compared to females [AOR = 1.99, 95% CI: 1.25–3.21].

Furthermore, respondents with good knowledge about hypertension were 2.82 times more likely to check their blood pressure compared to those with poor knowledge [AOR = 2.82, 95% CI: 1.73–4.61]. Those with a positive attitude towards hypertension prevention and control were 3.22 times more likely to engage in BP checkup practices than those with a negative attitude [AOR = 3.22, 95% CI: 1.99–5.22]. Additionally, participants with a family history of hypertension were significantly more likely to check their blood pressure, with an adjusted odds ratio of 4.86 [95% CI: 2.47–9.58].

## Discussion

Blood pressure checkup practices among federal ministry civil servants in this study were 43.8% [95% CI: 39.3% - 48.4%]. Age, sex, knowledge, attitude, and family history of hypertension were factors significantly associated with blood pressure checkup practice, having a p-value of less than 0.05.

This prevalence (43.8%) in this study was comparable to the findings of a study conducted in Nigeria, which reported a prevalence of 42.9% ^[48]^. However, it is notably higher compared to findings from studies conducted in other regions. For instance, a study in Ghana showed only 13.7% of individuals engaging in blood pressure checkups, which may reflect differences in healthcare infrastructure or cultural practices regarding preventive health measures ^[46]^. Similarly, in Bangladesh, 31.7% of people reported checking their blood pressure, which is still significantly lower than the rate observed among the federal ministry civil servants in this study ^[4]^. In Arba Minch Town, Ethiopia, only 29.8% of participants checked their blood pressure, indicating a lower level of health awareness or access ^[17]^. These variations highlight the importance of regional and contextual factors influencing health behaviors such as blood pressure checking.

The prevalence of blood pressure checkup practices among federal ministry civil servants is significantly lower compared to a study conducted among the Nepalese population, where 81.5% reported having blood pressure checkup practice ^[31]^. The remarkably higher rate in Nepal could be due to different factors, such as stronger health policies that promote routine health checks, more widespread community health programs, or cultural practices that encourage preventive health. These discrepancies point to a need for targeted interventions among federal civil servants to increase awareness and encourage more blood pressure checkups.

Furthermore, the blood pressure checkup practice in this study (43.8%) is lower compared to findings from other studies conducted in different populations. For instance, a study done in the general population of Ethiopia, the Addis Ababa report showed a higher blood pressure checkup practice of 59.8% ^[21]^. This difference suggests that even within the same city, there can be considerable variations in health-seeking behaviors among different groups.

The higher rate in the general population might be attributed to increased public awareness initiatives targeting the general public, or possibly greater utilization of health services among non-civil servants. This indicates that civil servants, despite relatively good knowledge and potential access to health services, may not be as proactive in engaging in blood pressure checkup practice.

This study, along with others, found that increasing age was a significant factor influencing blood pressure checkup practices. Federal ministry civil servants aged 40 years and above were 2.95 times more likely to participate in blood pressure checkups compared to those under 40 years old. Similarly, studies conducted in Arba Minch town, Ethiopia, and Nepal found that civil servants aged 40 and above were 5.75 and 1.6 times more likely to check their blood pressure, respectively ^[17,31]^. This difference could be attributed to increased awareness and concern about health risks that tend to grow with age. As people get older, they are generally more aware that age is a significant risk factor for hypertension, prompting them to be more proactive in checking their blood pressure. In contrast, younger civil servants may not perceive themselves to be at high risk for hypertension and might therefore be less inclined to engage in routine blood pressure checkup practice.

Gender differences regarding blood pressure checkup practices are significant, with males generally having higher rates of blood pressure checkup practice ^[31,49,50]^. In this study, male participants have 1.99 times higher blood pressure checkup practice as compared to females, which is similar to the study done in Nepal, where males are approximately 1.99 times more likely than females to participate in blood pressure checkup practices ^[31]^. This difference suggests that women may face barriers that limit their engagement in preventive healthcare, such as caregiving responsibilities and a lack of awareness about the importance of blood pressure checkups ^[50]^.

The study findings showed that hypertension knowledge was significantly associated with blood pressure checkup practice. In this study, civil servants with good knowledge of hypertension were found to be 2.82 times more likely to engage in blood pressure checkup practices as compared to those with poor knowledge.

Those who have a positive attitude towards the hypertension prevention mechanism have about 3.2 times higher blood pressure checkup practice compared with those who have a negative attitude. This might be because of their good knowledge and positive attitude towards hypertension prevention mechanisms, civil servants worry about chronic illnesses, and concerns about their health.

In this study, there is a significant association between family history of hypertension and the practice of blood pressure checkups among civil servants. Specifically, civil servants having a family history of hypertension were 4.9 times more likely to engage in a blood pressure checkup compared to those without a family history. The findings of this study align with and strongly support results from a similar study conducted in Arba Minch Town, which reported that civil servants with a family history of hypertension were 1.69 times more likely to check their blood pressure than those without a family history ^[17]^. Although the association is significant in both studies, the higher likelihood ratio in this study suggests that the study population in the federal ministry may have better access to healthcare resources, greater health literacy, or awareness of familial health risks may be more pronounced in the federal ministry context compared to the Arba Minch Town setting.

## Conclusion

Despite the importance of blood pressure checkups, a significant proportion of federal ministry civil servants in Addis Ababa do not engage in this practice, indicating a gap in health-seeking behavior related to hypertension prevention and control. Blood pressure checkup practice of civil servants was significantly related to factors such as age, Sex, knowledge, attitude, and family history of hypertension. These findings indicate the importance of targeted interventions to improve awareness, accessibility, and practices related to blood pressure checkups.

Strategies such as education campaigns focusing on younger individuals, those without a family history of hypertension, and females improve checkup practice. And efforts to improve knowledge and attitudes towards hypertension could significantly enhance blood pressure checkup practices. By implementing such measures, we can encourage more civil servants to prioritize their health and take proactive steps to prevent hypertension-related complications.

## Recommendations

Based on our study findings, it is recommended that the Ministry of Health and other relevant governmental and workplace stakeholders strengthen blood pressure screening services, particularly through the integration of routine checkups into workplace health programs.

There is a need to promote awareness campaigns focused on improving knowledge and attitudes towards hypertension prevention among civil servants. Health education efforts should be tailored to encourage lifestyle modifications and proactive screening behaviors, especially targeting younger employees and female staff, who are less likely to engage in Blood pressure checkup practices. Additionally, workplaces should consider implementing or expanding on-site health services, including regular BP monitoring, to enhance early detection and timely management of hypertension.

Researchers and program planners are encouraged to further investigate barriers to screening uptake and develop context-specific interventions. These coordinated efforts can help reduce the burden of undiagnosed hypertension and its associated complications among Ethiopia’s federal workforce.

## Data Availability

The data supporting the findings of this study will be made publicly available in Figshare upon acceptance of the manuscript.

## Limitations of the Study

Participants might recall their blood pressure checkup practices inaccurately, affecting data reliability. There were unmeasured behavioral variables not included in this study that might be influencing blood pressure checkup practices, like BMI, drinking Alcohol, and chewing khat.

## Acknowledgements

First of all, I would like to acknowledge Gamby Medical and Business College for providing technical assistance. Next, I want to express my profound gratitude to my advisor, Mr. Zewdu Minwuyelet, for his helpful and intellectual comments. Then, my beloved wife, Yenework Addis, I would like to thank you for supporting me despite you are in overwhelming challenges. Finally, I would like to thank the Ethiopian Federal Ministries’ administrative leaders and civil servants for their invaluable support during information sharing.

## Declaration

### Consent for Publication

Consent for publication was not applicable as this study does not contain identifiable patient data.

### Availability of Data and Materials

The datasets generated and/or analyzed during the current study are available from the corresponding author upon reasonable request. All relevant data supporting the findings of this study are included within the manuscript. Additional information can be provided to qualified researchers for academic and non-commercial purposes, in compliance with ethical and institutional regulations.

### Authors’ Contribution

Nibretie Chalachew Anteneh conceived and designed the study, collected and analyzed the data, interpreted the findings, and drafted the manuscript. Tadesse Shiferaw Chekol contributed to the development of the study design, supported data analysis and interpretation, and critically reviewed the manuscript for intellectual content. Zewudu Minwuyelet provided overall supervision, technical guidance, and advisory support throughout the research process and manuscript preparation. All authors read and approved the final version of the manuscript for submission.

## Other elements

### Human Participants Research Checklist

***Complete the following if your study involved human participants or human participants’ data. These questions should be addressed for prospective and retrospective studies***.

1. Did you obtain ethics approval for this study?
  - If yes, please upload (file type “Other”) all the approval documents you received from your ethics committee to cover the entire range of the study period (i.e. the original approval document and any extension documents). Where ethics approval was obtained from more than one study location, please provide approval document(s) from all of the sites. If the original document is in another language, please also provide an English translation. <u>✓</u> Uploaded –N/A
  - If you did not obtain ethical approval, please explain why this was not required below.
2. If you prospectively recruited human participants for the study – for example, you conducted a clinical trial, distributed questionnaires, or obtained tissues, data or samples for the purposes of this study, please report in the Methods:
  i. The day, month and year of the **start and end** of the recruitment period for this study.
  ii. Whether participants provided informed consent, and if so, what type was obtained (for instance, written or verbal, and if verbal, how it was documented and witnessed). If your study included minors, state whether you obtained consent from parents or guardians. If the need for consent was waived by the ethics committee, please include this information. Please state the line number(s) in the Methods where this is reported on **line two of subsection of study design in Methods section** <u>✓</u> Completed –N/A
3. If you are reporting a retrospective study of, for example, medical records, archived samples, survey data, please report in the Methods section:
  I. the day, month and year when the data were accessed for research purposes
  II. whether authors had access to information that could identify individual participants during or after data collection
  Please state the line number(s) in the Methods where this is reported Completed <u>✓</u>– N/A

---

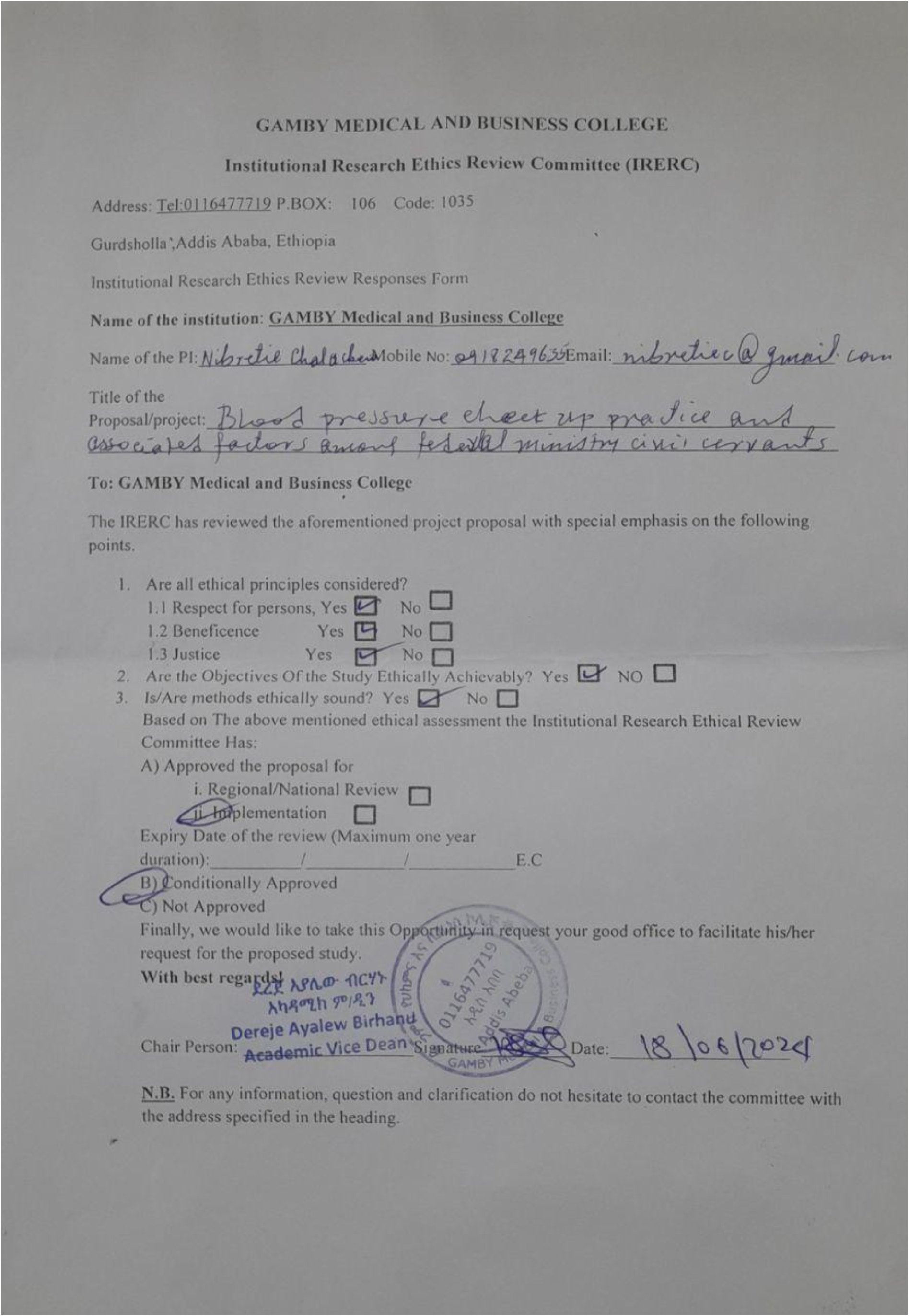

## References

1. Lewis MN, Shatat IF, Phillips SM. Screening for hypertension in children and adolescents: Methodology and current practice recommendations. Vol. 5, Frontiers in Pediatrics. Frontiers Media S.A.; 2017.

2. Mekonene M, Baye K, Gebremedhin S. Epidemiology of hypertension among adults in Addis Ababa, Ethiopia. Prev Med Reports. 2023 Apr 1;32.

3. Abebe SM, Berhane Y, Worku A, Getachew A. Prevalence and associated factors of hypertension: A cross-sectional community-based study in Northwest Ethiopia. PLoS One. 2015 Apr 24;10(4).

4. Rahman N, Alam SS. Knowledge, attitude, and practice about hypertension among adult people of selected areas of Bangladesh. MOJ Public Health. 2018;7(4):211–4.

5. Angaw K, Dadi AF, Alene KA. Prevalence of hypertension among federal ministry civil servants in Addis Ababa, Ethiopia: A call for a workplace-screening program. BMC Cardiovasc Disord [Internet]. 2015;15(1):1–6. Available from: http://dx.doi.org/10.1186/s12872-015-0062-9

8. Health M of E. National Strategic Plan For The Prevention And Control Of Major Non-Communicable Diseases. 2013.

9. Getahun GK, Arega M, Keleb G, Shiferaw A, Bezabih D. Assessment of routine medical checkups for common noncommunicable diseases and associated factors among healthcare professionals in Addis Ababa, Ethiopia, in 2022 a cross-sectional study. Ann Med Surg. 2023 May 1;85(5):1633–41.

10. Fazal F, Shahani HA. Attitudes and Factors Determining the Practice of Routine Medical Checkups in the People of Rawalpindi, Pakistan : A Cross-Sectional Study : 2023;(May):1–8.

11. Perspectives S. Is the Periodic Health Examination Worthwhile? Some Perspectives. Can Med Assoc. 1979;121:1193–254.

12. Aremu A.B.1, Afolabi I.B2, Salaam M1. Awunor N.S1, Sulayman A.A1, Ilori O1. NKU and SR. PREVALENCE AND FACTORS ASSOCIATED WITH ROUTINE MEDICAL CHECKUP AMONG PATIENTS ATTENDING MASAKA REGIONAL REFERRAL HOSPITAL, UGANDA Aremu. Int J Adv Res. 2021;9(07):97–105.

13. David T. Liss, PhD1; Toshiko Uchida M etal. General Health Checks in Adult Primary Care: A Review. MedRxiv. 2021;1(165):1–35.

14. Boulware LE, Marinopoulos S, Phillips KA, Hwang CW. Systematic Review: The Value of the Periodic Health Evaluation. Ann Intern Med. 2007;

15. Virgini V, Meindl-Fridez C, Battegay E, Zimmerli L. Check-up examination: Recommendations in adults. Vol. 145, Swiss Medical Weekly. EMH Schweizerischer Arzteverlag AG; 2015.

16. Christensen B, Lauritzen T. General health screenings to improve cardiovascular risk profiles : A randomized controlled trial in general practice with 5-year follow-up. J Fam Pract. 2002;51(6)(July):546–52.

17. Legisso TZ, Mamo BG, Bimrew AM, Fikadu T. Blood Pressure Examination Habit and Its Determinants Among Civil Servants in Arba Minch Town: A Cross-Sectional Study – Using Hurdle Poisson Regression Model. Integr Blood Press Control. 2023;16:1–9.

19. Noncommunicable N, Management D. Protocols National. National Management Protocol Noncommunicable Diseases. 2021.

20. Bayray A, Meles KG, Sibhatu Y. Magnitude and risk factors for hypertension among public servants in Tigray, Ethiopia: A cross-sectional study. PLoS One. 2018 Oct 1;13(10).

21. Bekele A, Gelibo T, Amenu K, Getachew T, Defar A, Teklie H, et al. The hidden magnitude of raised blood pressure and elevated blood glucose in Ethiopia: A call for initiating community based NCDs risk factors screening program. Ethiop J Heal Dev. 2017;31(Specialissue1):362–9.

6. Tiruneh SA, Bukayaw YA, Yigizaw ST, Angaw DA. Prevalence of hypertension and its determinants in Ethiopia: A systematic review and meta-analysis. Vol. 15, Public Library of Science. Public Library of Science; 2020.

7. Anker D, Santos-Eggimann B, Santschi V, Del Giovane C, Wolfson C, Streit S, et al. Screening and treatment of hypertension in older adults: Less is more? Vol. 39, Public Health Reviews. BioMed Central Ltd.; 2018.

18. Siu AL, Services USP, Force T. Screening for High Blood Pressure in Adults : U. S. Preventive Services Task Force Recommendation Statement. Ann Intern Med. 2015;163(10):778–86.

22. Roba HS, Beyene AS, Mengesha MM, Ayele BH. Prevalence of Hypertension and Associated Factors in Dire Dawa City, Eastern Ethiopia: A Community-Based Cross-Sectional Study. Int J Hypertens. 2019;2019.

23. Alzahrani AMA, Felix HC, Stewart MK, Selig JP, Swindle T, Abdeldayem M. Utilization of Routine Medical Checkup and Factors Influencing Use of Routine Medical Checkup among Saudi Students Studying in the USA in 2019. Saudi J Heal Syst Res. 2021;1(1):16–25.

24. Bayor S. Periodic Medical Checkup Among Health Workers at a Teaching Hospital in Ghana. Res Sq [Internet]. 2023; Available from: 10.21203/rs.3.rs-2533456/v1

25. AL-Kahil AB, Khawaja RA, Kadri AY, Abbarh, MBBS SM, Alakhras JT, Jaganathan PP. Knowledge and Practices Toward Routine Medical Checkup Among Middle-Aged and Elderly People of Riyadh. J Patient Exp. 2020 Dec;7(6):1310–5.

26. Du M, Li P, Tang L, Xu M, Chen X, Long H. Cognition, attitude, practice toward health checkup and associated factors among urban residents in southwest China, Sichuan province, 2022: a community-based study. J Public Heal. 2023;

27. San TS, Plianbangchang S. Knowledge, attitude and practice of preventive behavior toward hypertension among Myanmar migrants in Samut Sakhon province, Thailand. J Heal Res. 2018;32(1):32.

28. Article O. Factors Affecting Routine Medical Screening Among Health Workers in a Tertiary Hospital in Delta State. Abadom EG, Otene. 2022;951–9.

29. Noguchi R. Factors affecting participation in health checkups : Evidence from Japanese survey data. Health Policy (New York). 2019;123(4),:360–366.

30. Yilmaz S, Calikoglu EO, Kosan Z. Factors Affecting Routine Medical Screening Among Health Workers in a Tertiary Hospital in Delta State. Niger J Clin Pract. 2019;22(July):1070–7.

31. Gupta R Das, Haider SS, Sutradhar I, Hasan M, Joshi H, Haider MR, et al. Gender differences in hypertension awareness, antihypertensive use and blood pressure control in Nepalese adults: findings from a nationwide cross-sectional survey. J Biosoc Sci. 2020;52(3):412–38.

32. Ababa A, Achamo T. Utilization of Preventive Medical Check-Up and Associated Factors Among Public Hospital Workers. 2024;1–24.

33. Maulana AE. Regular medical checkup behaviour : preventing is better than curing. Asia Pacific J Mark Logist. 2017;30(2):478–94.

34. Kohori H, Id S, Uematsu H, Id ND, Wangdi U, Dorjee C, et al. Social and behavioral factors related to blood pressure measurement: A cross-sectional study in Bhutan. PLoS ONE, [Internet]. 2022;17(8 August):1–17. Available from: http://dx.doi.org/10.1371/journal.pone.0271914

35. Leigh B, Guisinger D, Fech J. Blood Pressure Screening in the Workplace. AAOHN J. 1989;37(1):14–17.

36. Mozu IE, Marfo AFA, Opare Addo M, Buabeng KO, Owusu-Daaku FT. Evaluation of workplace hypertension preventative and detection service in a Ghanaian University. Int J Pharm Pract. 2023;31(2):237–42.

37. Weldeghebrael EH. Addis Ababa: City Scoping Study. African cities Res consertium. 2021;(June):1–10.

38. Central Statistics Agency. Addis Ababa, Ethiopia Metro Area Population 1950-2021. 2021;1–4.

39. Ayalew TL, Wale BG, Zewudie BT. Burden of undiagnosed hypertension and associated factors among adult populations in Wolaita Sodo Town, Wolaita Zone, Southern Ethiopia. BMC Cardiovasc Disord [Internet]. 2022;22(1):1–9. Available from: https://doi.org/10.1186/s12872-022-02733-3

40. Guda T. Assessment of Knowledge, Attitude and Practice Towards Prevention and Control Of Hypertension Among Members Of The Ethiopian Army Assigned For Peace Keeping Mission. Jouna. 2015;(June).

41. Mahmud Ahmed S, Belaye Teferi M. Assessment of Knowledge, Self-care Practice, and Associated Factors Among Hypertensive Patients the Public Hospital of Addis Ababa Ethiopia 2016 G.C. Int J Cardiovasc Thorac Surg. 2020;6(2):28.

42. Federal Ministry of Health of Ethiopia. Guidelines on Clinical and Programmatic Management of Major Non-Communicable Diseases. 2016.

43. Keller K. National Center for Health Statistics. Encycl Obesity SAGE Publ Inc. 2014;1–2.

44. Dhungana RR, Karki KB, Bista B, Pandey AR, Dhimal M, Maskey MK. Prevalence, pattern and determinants of chronic disease multimorbidity in Nepal: Secondary analysis of a national survey. BMJ Open. 2021;11(7):2020–1.

45. Han H ra, Chan K, Song H, Nguyen T, Mph MSN. Development and Evaluation of a Hypertension Knowledge Test for Korean Hypertensive Patients. J Clin Hypertens. 2011;13(10):750–7.

46. Addo J, Smeeth L. Prevalence, detection, management, and control of hypertension in Ghanaian civil servants Article. Glob Heal. 2016;(September 2008).

47. Working S, Town M, Kalssa A, Ayele G, Tamiso A, Girum T. Prevalence and Associated Factors of Hypertension among Civil. 2016;5(4):375–83.

48. Azubuike S, Kurmi R. Awareness, practices, and prevalence of hypertension among rural Nigerian women. Arch Med Heal Sci. 2014;2(1):23.

49. Patel R, Chauhan S. Gender differential in health care utilisation in India. Clin Epidemiol Glob Heal [Internet]. 2020;8(2):526–30. Available from: https://doi.org/10.1016/j.cegh.2019.11.007

50. Bhandari B, Narasimhan P, Vaidya A, Subedi M, Jayasuriya R. Barriers and facilitators for treatment and control of high blood pressure among hypertensive patients in Kathmandu, Nepal: a qualitative study informed by COM-B model of behavior change. BMC Public Health. 2021;21(1):1–14.

